# Prenatal Exposure to Emerging Pesticides and Childhood Allergy Risk: A First Mixture Assessment in an Urban Cohort

**DOI:** 10.1101/2025.11.13.25340136

**Authors:** Sergio Gómez-Olarte, Stefan Röder, Michael Borte, Martin Krauss, Werner Brack, Ana C. Zenclussen, Gunda Herberth, Carolin Huber

**Author notes:** **Corresponding Authors** Gunda Herberth, PhD. Department of Environmental Immunology, Helmholtz Centre for Environmental Research – UFZ, Permoserstraße 15, 04318, Leipzig, Germany., Sergio Gómez-Olarte, PhD. Department of Environmental Immunology, Helmholtz Centre for Environmental Research – UFZ, Permoserstraße 15, 04318, Leipzig, Germany. Shared last authorship (Gunda Herberth, Carolin Huber).

## Abstract

Pesticide gestational exposure may contribute to the development of allergies in childhood, yet evidence on its health impact on urban populations remains limited. This study investigates the association between prenatal exposure to individual and mixed pesticides and allergic outcomes, including asthma, wheezing, and eczema, at age 6 in 387 mother-child pairs from the LiNA cohort. Forty pesticides and metabolites were detected in urine during pregnancy through non-targeted screening, and 11 were selected (detection rate≥17%) for further analysis. Multivariable logistic regression models adjusted for covariates revealed statistically significant associations between dihydroxy-pyrimethanil and asthma (aOR=1.36, 95% CI: 1.04–1.80), and fluazifop-desbuthyl and wheezing (aOR=1.15, 95% CI: 1.01–1.31). No significant associations were observed for eczema. The mixture effect analysis with weighted quantile sum (WQS) regression showed that higher pesticide co-exposures significantly increased wheezing odds (aOR=2.08, 95% CI: 1.21–3.56). The main components of the WQS index were fluazifop-desbuthyl, flonicamid, hydroxy-metazachlor, and terbuthylazine, accounting for 67% of the overall dose-additive effect. These findings suggest that prenatal exposure to pesticides at dietary levels may increase the risk of childhood asthma and wheezing. Further epidemiological studies should replicate our findings by considering exposures to other pesticides of concern and their metabolites.

**Synopsis:** Risk assessment of chemical mixtures should cover dietary exposure to non-persistent pesticides in urban populations by screening for compound-specific metabolites.

## 1. Introduction

Some of the pesticides introduced during the Green Revolution are listed as persistent organic pollutants, including dichlorodiphenyltrichloroethane (DDT) and hexachlorobenzene.^1^ However, they were phased out by the 2000s due to growing public health concerns, particularly for pregnant women and children.^2^ For instance, a meta-analysis of seven European population cohorts showed that prenatal exposure to DDE, the main DDT metabolite, and PCB153 was associated with bronchitis and wheezing in children aged 4 years old.^3^ This suggests that gestation is a key window of susceptibility to pesticide exposure, as the fetal immune system is still in the developing phase.^4^

Gradually, persistent pesticides have been replaced with less persistent target-specific compounds, such as carbamates and neonicotinoids.^5^ Despite modern pesticides displaying an abridged human toxicity, scarce evidence indicates they could be associated with the development of allergic disorders early in life.^6^ In particular, the Costa Rican ISA cohort reported an association of current, but not prenatal, levels of pyrethroid metabolites with wheezing, itchy skin rash, and asthma in 5-year-old children.^7, 8^ This aligns with the findings of a French pilot study on the association between current exposure to ethylene thiourea, a mancozeb metabolite, and asthma and rhinitis at ages 3-10 years.^9^

Most available epidemiological evidence about pesticide effects on allergic outcomes derives from populations living in rural farming areas, where maternal exposure is at least five times higher than in the general public.^10^ Therefore, it does not reflect the health risks in urban environments, in which pesticide exposure primarily occurs through diet at lower doses.^11^ In this regard, a New York cohort found that prenatal exposure to the insecticide permethrin via indoor air increased the odds of cough, but not asthma or wheezing, in children 5-6 years old.^12^ To date, only a French longitudinal study has explored how urban pesticide co-exposures during pregnancy might contribute to childhood allergies,^13^ yet it overlooked mixture interactions that might enhance toxic effects.

The present study aims to investigate whether prenatal exposure to pesticides and their metabolites, detected by non-targeted analysis, is linked with the development of asthma, wheezing, and eczema in preschool children from the LiNA cohort. We addressed, for the first time, the health effects of single and mixed pesticides with a focus on the potential dose-additive effects of co-exposures.

## 2. Materials and Methods

### 2.1. Study design and recruitment

The Lifestyle and environmental factors and their influence on the Newborn Allergy risk - LiNA prospective cohort includes 622 pregnant women recruited between 2006 and 2008 in Leipzig, Germany. The study was approved by the Review Board of Leipzig University (file reference no. 046-2006). All participants voluntarily signed the informed consent. This study comprises 581 mothers who provided urine samples and questionnaire information at gestational week (WG) 34-36 and their paired singleton children aged 6 years. Statistical analyses were performed in 387 mother-child pairs with exposure assessments and complete socio-demographic data, including occupation (**Fig. S1**).

### 2.2. Chemical exposure assessment

Pesticides and their metabolites were detected in the first morning void urine samples, which were collected and stored in polypropylene tubes at -80°C until analysis. After thawing, 500 µL aliquots were used for sample preparation and measurement using liquid chromatography-high-resolution mass spectrometry (LC-HRMS), as previously described.^14^ The following pesticides or corresponding metabolites detected in at least n=100/581 mothers (detection rate: DR≥17 %) were selected for the health assessment: metalaxyl, carbetamide, terbuthylazine, imidacloprid, flonicamid, hydroxy-isoproturon, dihydroxy-pyrimethanil, hydroxy-simazine, hydroxy-propamocarb, fluazifop-desbuthyl, and hydroxy-metazachlor. The non-targeted measurements provided uncorrected peak intensities, referred to as levels onwards, but not precise chemical concentrations. When more than one marker of a pesticide was available, the parent compound or metabolite with the highest DR measurement was implemented in the regression models.

### 2.3. Allergic outcomes and covariates

Information on childhood allergies and related symptoms (asthma, wheezing, and eczema) was collected via annual self-administered questionnaires, asking the parents for a doctor’s diagnosis in the previous 12 months. Yearly allergy reports were used to define the lifetime prevalence at year 6 for each outcome. Besides, general socio-demographic and lifestyle factors were retrieved from medical records and questionnaires applied during the third trimester of pregnancy and annually after the child’s birth. Among these variables, smoking or environmental tobacco smoke (ETS) exposure during pregnancy, breastfeeding throughout the first 6 months, family history of atopy, parental education level, and child sex were identified as potential covariates based on previous literature^15^ (see directed acyclic graph in **Fig. S2**).

### 2.4. Statistical analysis

Given right-skewed distributions, chemical peak intensities were log_2_-transformed for regression analysis and presented using the geometric mean (GM) and percentiles. Mother-child pairs’ characteristics and covariates were described with numbers (n) and percentages (%) and compared among pregnancy and 6-year follow-up observations using a Chi-square or Fisher’s exact test (n<5). Statistical significance was defined with a *p*-value threshold of 0.05.

Individual associations between 11 pesticides detected in the mothers (DR≥17%) and the children’s outcomes (asthma, wheezing, and eczema) were investigated using multivariable logistic regression models, crude and adjusted for covariates. Exposure values below the detection limit (BDL) were replaced by the mean of 20 imputations generated via a truncated log-normal function as recommended before^16^, setting the lowest measured peak intensity value (limit of detection: LOD) divided by √2 as the compound-specific detection threshold. The dataset resulting from this multiple imputation (MI) was then log_2_-transformed to fit the single regression models. Another exposure dataset was created by imputing BDL values with LOD/√2 and used for sensitivity analysis of the multivariable regression estimates.

Furthermore, multi-pollutant analyses accounting for potential dose-additive effects among the mixture components were conducted using weighted quantile sum (WQS) regression.^17^ Pesticide and metabolite measurements were categorized into 3 quantiles (Qs); values BDL were assigned to Q1 (non-exposed/low exposure), while those detected were scored to Q2 and Q3 (medium and high exposures). This strategy offers the best model fit, accuracy, and statistical power to identify chemicals contributing to mixture effects when exposure values BDL are 50-80%^18^, as for our pesticide exposure data. When fitting the crude and adjusted WQS with each allergy outcome, the exposure data were randomly split into training and validation sets (0.40 and 0.60) to estimate single chemical weights by the bootstrap method (n=200) and the repeated holdout validation procedure (n=20).^19^ Pesticides with weights >9% were identified as the main driver of the mixture effects, based on the cut-off τ value (inverse of exposure, n=11).^17^

To assess exposure risk, non-adjusted and adjusted odds ratios (OR/aOR) and 95% confidence intervals (CIs) were estimated from logistic regression coefficients, with values >1.0 indicating increased risk. Statistical analyses and plots, including correlation matrices and WQS regression models, were computed with the R software (v4.4.1) using the packages “corrplot” (v0.95)^20^ and “gWQS” (v3.0.5).^21^

## 3. Results and Discussion

### 3.1. Sociodemographic characteristics, allergic outcomes, and pesticide exposure levels

The present study included a subgroup of 387 mother-child pairs, whose sociodemographic and lifestyle features were similar (*p*>0.05) to the entire LiNA cohort with pesticide exposure assessment during pregnancy (n=581). Among these variables, smoking/ETS exposure, parental education level, breastfeeding during the first 6 months, history of atopy, and child’s sex were included as covariates in all regression models (**Table S1**). The lifetime prevalence of asthma, wheezing, and eczema in the 6-year-old children was 4.9%, 43%, and 22%, respectively. Other studies on pesticide exposure have reported a higher prevalence of asthma (7.7% and 12%) and a similar or lower prevalence of wheezing (40% and 32%) in 5-year-old children.^7, 13^ Compared to the LiNA cohort, the prevalence of eczema in children from these studies (7% and 48%) was markedly different.^7, 13^

Non-targeted analyses revealed that children were prenatally exposed to mixtures of pesticides and their derived metabolites, which are commonly used as fungicides, herbicides, and insecticides (**Fig. 1A**). The mothers did not report occupational pesticide exposure; thus, dietary intake was the most likely chemical source, as demonstrated by our analysis of polyphenol food markers.^14^ The distribution levels of pesticides and metabolites are shown in **Table S2**. Human pesticide co-exposure is linked to common sources like food, resulting in exposure matrices with correlated components.^22^ Thus, we explored the collinearity between the pesticides and metabolites via Spearman’s rank correlation matrices of the original, MI-imputed, and LOD/√2-imputed datasets. Findings were consistent across the three exposure matrices (**Fig. 1B**/**Fig. S3**), with positive (*r*≥0.65; *p*<0.05) correlations between metalaxyl and carbetamide, carbetamide and hydroxy-isoproturon, and hydroxy-isoproturon and hydroxy-metazachlor. Since it has been shown that assigning truncated random MI values to measures BDL generates unbiased regression parameter estimates^16^, we employed the log_2_-transformed MI-imputed dataset to fit the multivariable regression models.

**Figure 1.**
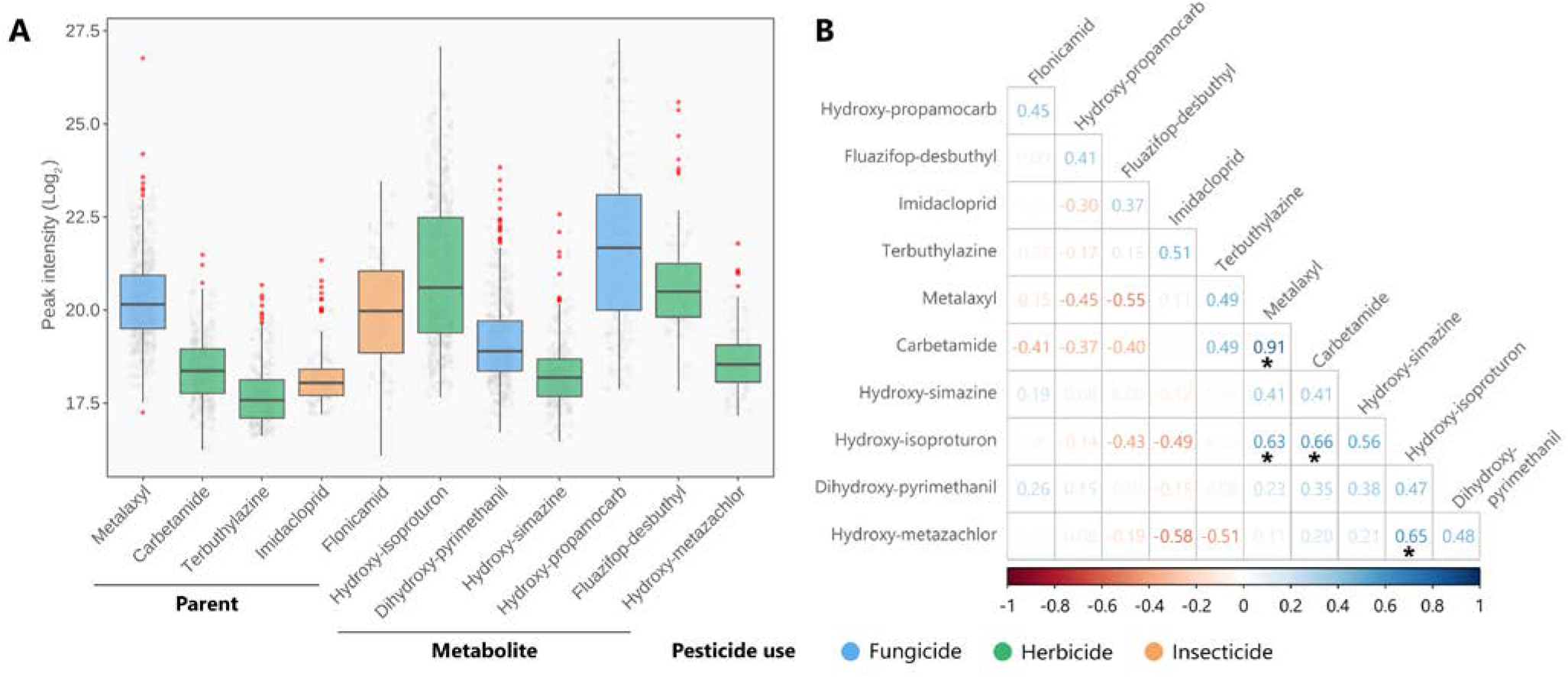
(**A**) Box plot that displays the distribution of pesticides and metabolites measured (log_2_-transformed) in urine during pregnancy (WG 34-36); red dots denote extreme values. Chemicals are shown from left to right according to their type (parent or metabolite) and DR (highest to lowest). (**B**) Pairwise Spearman’s rank correlation matrix of chemical exposure levels, where values BDL were replaced with MI means (n=20). In the color spectrum, blue and red shades show positive and negative correlations between the chemicals. The asterisk (*) denotes statistically significant correlations (*p*<0.05).

### 3.2. Association of pesticides and metabolites with allergic outcomes

We investigated the association between the individual pesticides (parent, n=5; metabolites, n=6) and allergic outcomes, including asthma, wheezing, and eczema, in 6-year-old children, by multivariable logistic models after controlling for covariates (**Fig. S2**). The precise ORs, 95% CI, and *p*-values for all estimates are depicted in **Table S3**. Both the crude and adjusted models showed a statistically significant positive association of dihydroxy-pyrimethanil with asthma (aOR=1.36, 95% CI: 1.04-1.80) and fluazifop-desbuthyl with wheezing (aOR=1.15, 95% CI: 1.01-1.31); whereas, there was no relationship between the chemical exposures and eczema (**Fig. 2**). Thus, prenatal exposure to doubling levels of pyrimethanil and fluazifop metabolites increased the odds of asthma and wheezing by 36% and 15% in children 6 years old, respectively. Sensitivity analysis with the LOD/√2-imputed exposure dataset consistently showed positive associations (*p*<0.05) and ORs of similar magnitude between dihydroxy-pyrimethanil and asthma, and fluazifop-desbuthyl and wheezing (**Table S4**).

**Figure 2.**
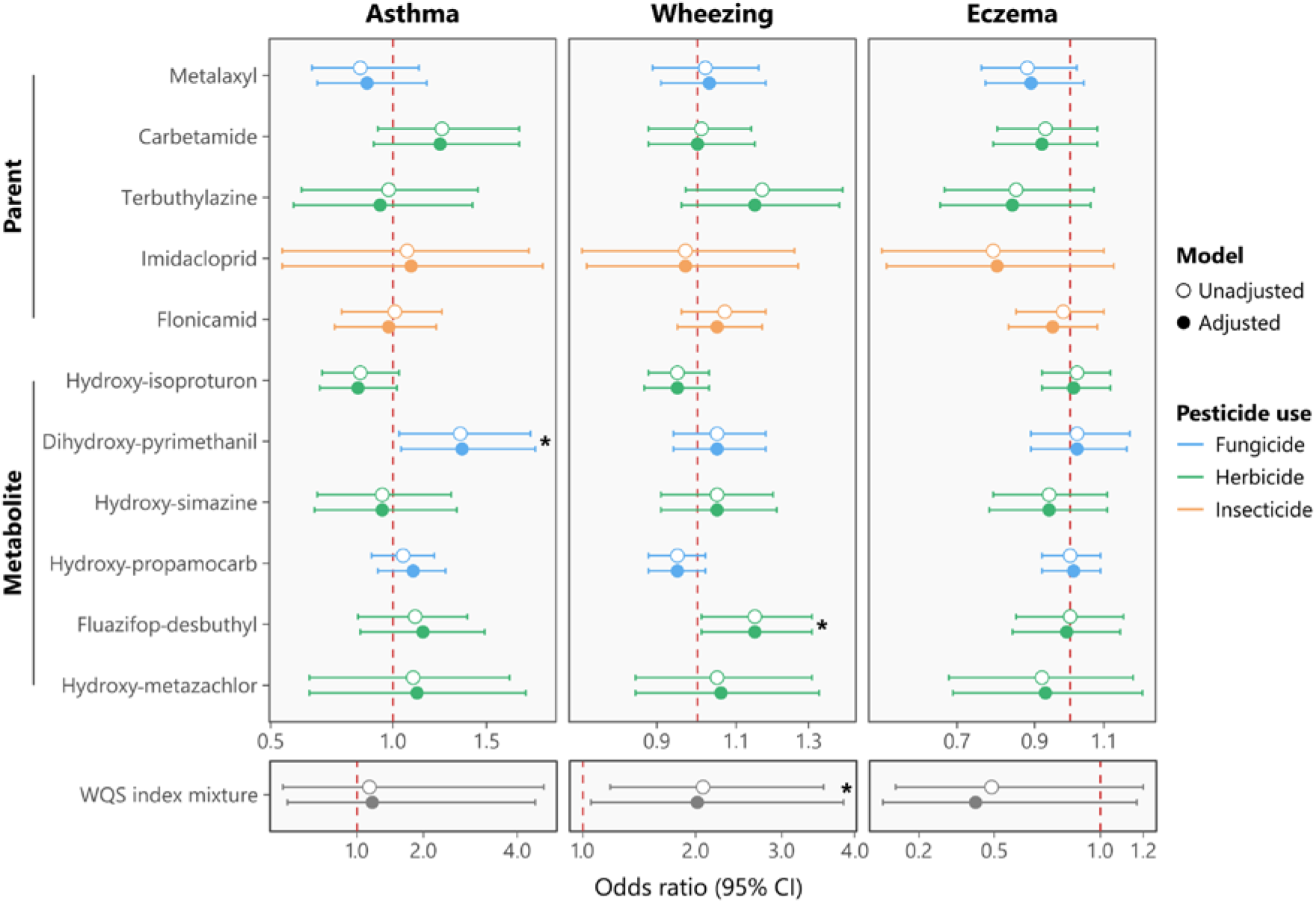
Odds ratio plot of the association between prenatal exposure to 11 pesticides (parent and metabolite compounds) and the main allergic outcomes, asthma, wheezing, and eczema, in 6-year-old children (n=387). The logistic regression models were adjusted for smoking/ETS exposure during pregnancy, breastfeeding up to 6 months, parental atopy history, parental education level, and child sex. Chemicals are shown top-down according to their type (parent or metabolite) and DR (highest to lowest). The precision of the estimates is indicated using 95% CIs. The asterisk (*) denotes statistically significant associations (*p*<0.05).

Although epidemiological evidence on the health impacts of pesticides is moderately consistent, only limited studies have examined prenatal exposures. In addition, comparisons across these findings are challenging due to differences in population exposure levels, detected chemical forms, and the timing of outcome assessment. For instance, the Costa Rican ISA cohort did not find a relationship between prenatal levels of hydroxy-pyrimethanil and wheezing during the first year of life,^8^ which aligns with our observation of no association between dihydroxy-pyrimethanil and wheezing at age 6. This showcase concordant results for an outcome using two different metabolites of pyrimethanil at infancy and preschool age; yet, we found that dihydroxy-pyrimethanil was linked to asthma. The LiNA urine samples that contained polyphenol markers for citrus fruit consumption featured higher DRs of pyrimethanil metabolites,^14^ suggesting this specific fruit type is a likely source of exposure.

Next, we assessed whether prenatal exposure to the pesticide mixture, containing parent and metabolite compounds, was associated with allergic outcomes in the 6-year-old children using WQS regression. The WQS models of asthma, wheezing, and eczema were adjusted for covariates and restricted to the positive index to examine potential dose-additive effects among the exposure matrix. The pesticide WQS index exhibited a statistically significant positive association (β=0.70, 95% CI: 0.06-1.34) with wheezing. This indicates that a one-quantile increase (from low to medium and medium to high exposure levels) in the WQS index doubled the odds of wheezing in children aged 6 years (aOR=2.08, 95% CI: 1.21-3.56). On the contrary, the relationships between the pesticide WQS index and asthma (dihydroxy-pyrimethanil weight=15%) and eczema were not statistically significant (**Fig. 2**). The main contributors to the overall mixture effect on wheezing were fluazifop-desbuthyl (26%), flonicamid (19%), hydroxy-metazachlor (11%), and terbuthylazine (11%), which had a combined weight of 67% (**Fig. 3**). As of now, no human toxicological studies have investigated the potential mechanisms linking prenatal exposure to these pesticides with the development of asthma and its related symptoms (e.g., wheezing). Nevertheless, fluazifop-butyl has been shown to induce oxidative stress and apoptosis in mouse testis cells *in vitro*,^23^ whereas terbuthylazine exposure triggers myocardial damage in chickens via activation of the cGAS– STING/NF-κB proinflammatory pathway.^24^ This evidence renders plausible the role of the identified pesticides of concern in promoting inflammation and narrowing of the airways through immune-related mechanisms.^25, 26^

**Figure 3.**
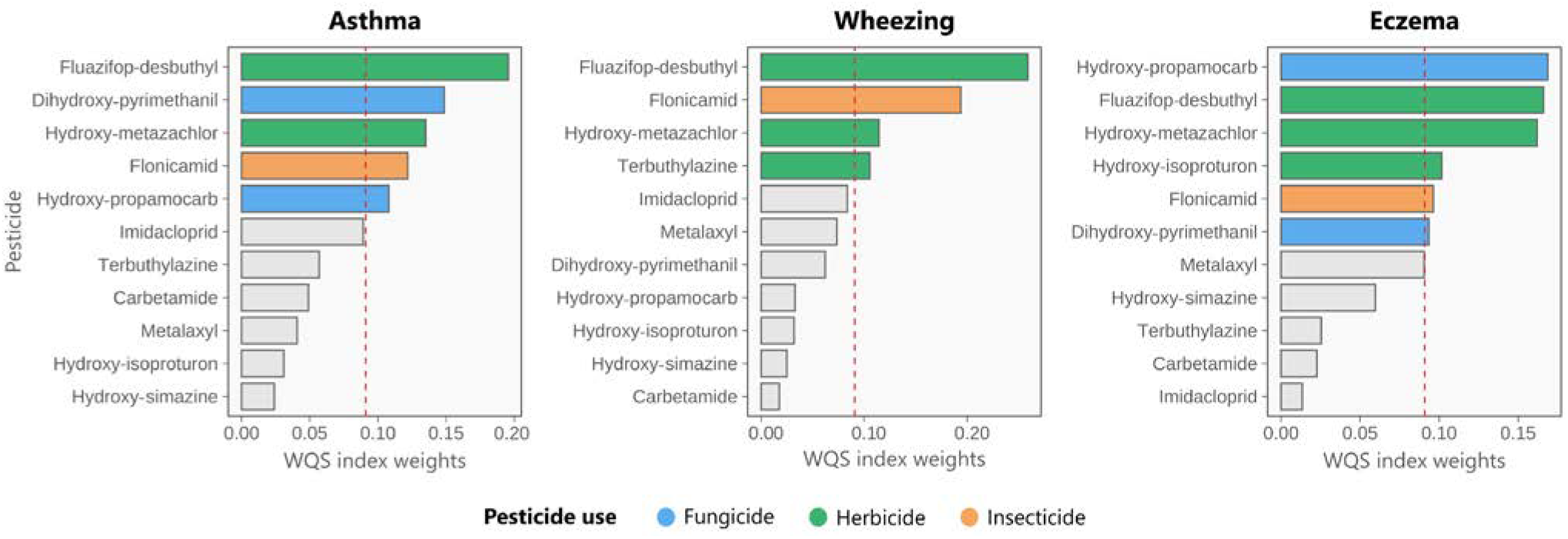
Weight magnitudes from the WQS index for the association of pesticides and metabolites with asthma, wheezing, and eczema prevalence in 6-year-old children. Estimates of the WQS regression were adjusted for smoking/ETS exposure during pregnancy, breastfeeding up to 6 months, parental atopy history, parental education level, and child sex. The dashed red lines represent the cut-off *τ* (inverse of exposure, n=11) to define the exposures driving the effect within the chemical mixture.

To our knowledge, only one recent study from the ELFE birth cohort has investigated pesticide mixture effects during early life. They reported an increased risk of wheezing in 5.5-year-old children when prenatally exposed to high levels of mixed trace elements and pesticides, containing pyrimethanil, metalaxyl, and imidacloprid, detected in the mother’s diet.^13^ Although the study featured a robust sample size, the mixture analysis relied on individual exposure scores derived from linking nutritional questionnaires to pesticide measurements in foods,which restricted the accuracy of the exposure assessment. Notably, the USA Green Housing multi-city cohort investigated the association between current exposure to a pesticide metabolite mixture and asthma outcomes in a small sample (n=162) of children ages 7-12 years and found no relationship.^27^ Therefore, the current work provides the first evidence on how dose-additive effects of prenatal pesticide co-exposures, at non-occupational levels, may contribute to developing childhood wheezing. Moreover, our findings suggest that routine chemical monitoring of food should not be restricted to parent compounds,^28^ but rather consider pesticide metabolites as markers of internal exposure, especially for the risk assessment of chemical mixtures. ^29^

### 3.3. Strengths and limitations

This study has several strengths. LiNA cohort’s longitudinal and prospective design enabled describing a likely causal association between prenatal exposure to a pesticide mixture and wheezing risk. By implementing appropriate WQS mixture modeling, we could identify the major chemicals, three herbicides and one insecticide, driving this dose-additive effect. The robustness of the regression estimates was tested by fitting crude and adjusted models with exposure datasets completed with MI and fixed (LOD/√2) values. Thus, in contrast to most studies, we considered exposures with about 20% of detected values for the health assessment and found they may contribute to chemical cocktail effects. In particular, it shows that metabolites can have higher DRs than parent compounds, making them more reliable for exposure assessment.^30^ Nonetheless, the present study also has limitations related to the exposure assessment and children’s outcome prevalence. As pesticides were measured in a unique random urine sample during pregnancy, we may have overlooked pesticides or other chemicals with short half-lives, such as carbamates, or residual confounding due to postnatal pesticide exposures. The non-targeted screening only provides signal intensity values that correlate with the concentration of the pesticides and metabolites. This limits our capability to compare exposure levels with those of other populations. Besides, the collection of allergic outcome information by self-report questionnaires has the potential for recall and misclassification biases. The rather low prevalence of asthma (<5%) in the LiNA cohort at age 6 may have restricted the statistical power of the regression models to detect a true association with the pesticide mixture. Therefore, we recommend replication studies in larger populations with comparable exposure patterns.

## Data Availability

All data produced in the present study are available upon reasonable request to the authors.

## Supporting Information

Results on the sociodemographic and lifestyle characteristics of LiNA cohort mother-child pairs (**Table S1**); distribution of pesticide and metabolite peak intensities in urine samples of LiNA mother-child pairs at age 6 (**Table S2**); output from logistic regression models for prenatal exposure to pesticides and allergies in 6-year-old children (n=387), using the MI-imputed (**Table S3**) and LOD/√2-imputed (**Table S4**) exposure datasets; flow chart of recruitment and inclusion criteria of LiNA mother-child pairs at age 6 (**Fig. S1**); directed acyclic graph that displays the causal pathway linking pesticide prenatal exposure to allergic outcomes (**Fig. S2**); and Spearman’s rank correlation matrices of chemical exposure levels for the original and LOD/√2-imputed datasets (**Fig. S3**) (DOCX).

## Authorship Contribution

SGO: Conceptualization, Formal analysis, Investigation, Methodology, Visualization, Writing – original draft, Writing – review & editing. SR: Data curation, Methodology. MB: Investigation, Resources. MK: Funding acquisition, Project administration. WB: Resources. ACZ: Funding acquisition, Project administration, Writing – review & editing. GH: Conceptualization, Supervision, Writing – review & editing. CH: Investigation, Validation, Writing – review & editing.

## Notes

The authors declare no competing financial interest.

## Acknowledgments

This is a study from the ENDOMIX project: Understanding how endocrine disruptors and chemical mixtures of concern target the immune system to trigger or perpetuate disease. ENDOMIX has received funding from the European Union’s European Health and Digital Executive Agency under grant agreement No. 101136566. Views and opinions expressed are however those of the author(s) only and do not necessarily reflect those of the European Union or the European Health and Digital Executive Agency. Neither the European Union nor the granting authority can be held responsible for them. The LiNA study was supported by intramural funding from the Department of Environmental Immunology at the Helmholtz Centre for Environmental Research – UFZ. We thank Melanie Bänsch, Maik Schilde, and Michaela Loschinski for their technical support, and the participants of the LiNA cohort for their engagement with the study.

## Notes

### Competing Interest Statement

The authors have declared no competing interest.

### Author Declarations

The Lifestyle and environmental factors and their influence on the Newborn Allergy risk (LiNA) prospective birth cohort recruited 622 pregnant women between 2006 and 2008 in Leipzig, Germany, under the approval of the Review Board of Leipzig University (file No. 0462006). All participants signed the informed consent voluntarily.

