## Supporting Information for "Prenatal Exposure to Emerging Pesticides and Childhood Allergy Risk: A First Mixture Assessment in an Urban Cohort"

[a] Department of Environmental Immunology, Helmholtz Centre for Environmental Research – UFZ, Leipzig, Germany
[b] St. Georg Hospital Leipzig, Department of Pediatrics, Leipzig, Germany
[c] Department of Exposure Science, Helmholtz Centre for Environmental Research – UFZ, Leipzig, Germany
[d] Department Evolutionary Ecology & Environmental Toxicology, Faculty of Biological Sciences – Goethe University Frankfurt, Frankfurt am Main, Germany
[e] German Center for Child and Adolescent Health (DZKJ), Partner Site Leipzig/Dresden, Germany
[f] Perinatal Immunology, Saxonian Incubator for Clinical Translation (SIKT), Medical Faculty, Leipzig University, Leipzig, Germany
[#] Shared last authorship

*** Corresponding Authors**

Gunda Herberth, PhD. Department of Environmental Immunology, Helmholtz Centre for Environmental Research – UFZ, Permoserstraße 15, 04318, Leipzig, Germany.
Sergio Gómez-Olarte, PhD. Department of Environmental Immunology, Helmholtz Centre for Environmental Research – UFZ, Permoserstraße 15, 04318, Leipzig, Germany.

### Supporting Tables

**Table S1.** Sociodemographic and lifestyle characteristics of LiNA cohort mother-child pairs with pesticide exposure assessment at baseline (pregnancy) and the 6-year follow-up.

| Characteristic, n (%) | Pregnancy [a] n = 581 | 6-year follow-up [b] n = 387 | *p*-value [c] |
|---|---|---|---|
| **Maternal age at delivery (years)** | | | 0.649 |
| < 25 | 61 (10%) | 34 (8.8%) | |
| 25-30 | 223 (38%) | 146 (38%) | |
| 30-35 | 197 (34%) | 130 (34%) | |
| > 35 | 100 (17%) | 77 (20%) | |
| **Smoking/ETS exposure during pregnancy** | | | 0.074 |
| No | 470 (83%) | 335 (87%) | |
| Yes | 98 (17%) | 50 (13%) | |
| Missing | 13 | 2 | |
| **Parental school education [d]** | | | 0.239 |
| Low | 16 (2.8%) | 5 (1.3%) | |
| Medium | 128 (22%) | 79 (20%) | |
| High | 437 (75%) | 303 (78%) | |
| **Breastfeeding (1-6 months)** | | | 0.263 |
| No | 121 (22%) | 73 (19%) | |
| Yes | 417 (78%) | 303 (81%) | |
| Missing | 43 | 11 | |
| **Family history of atopy** | | | 0.978 |
| None | 192 (33%) | 127 (33%) | |
| One parent | 274 (47%) | 185 (48%) | |
| Both parents | 115 (20%) | 75 (19%) | |
| **Child sex** | | | 0.537 |
| Male | 303 (52%) | 194 (50%) | |
| Female | 278 (48%) | 193 (50%) | |

[a] LiNA cohort mothers with measurements of pesticides and their metabolites in urine and questionnaire data, n = 581 out of 622.

[b] Mother-child pairs at year 6 with complete outcome data.

[c] The Chi-square or Fisher's exact test (n < 5).

[d] Parental education defined by schooling years: 9 or fewer years (low), 10 years (medium), and 12 or more years (high).

**Table S2.** Distribution of pesticide and metabolite peak intensities measured in urine samples of LINA cohort mother-child pairs at age 6 years.

| Compound | DR | GM | Min | Percentile | | | | | Max |
|---|---|---|---|---|---|---|---|---|---|
| | | | | 5th | 25th | 50th | 75th | 95th | |
| **Parent pesticide** | | | | | | | | | |
| Metalaxyl | 0.90 | 1270366 | 187610 | 402920 | 755529 | 1164006 | 2007717 | 5048888 | 12413750 |
| Carbetamide | 0.57 | 344496 | 110515 | 149474 | 227315 | 329036 | 484946 | 962120 | 2927008 |
| Terbuthylazine | 0.45 | 215611 | 100679 | 108648 | 139098 | 195271 | 280879 | 784875 | 1666377 |
| Imidacloprid | 0.20 | 288459 | 149600 | 155023 | 210011 | 264976 | 320968 | 827975 | 2643546 |
| Flonicamid | 0.17 | 1086172 | 69116 | 190997 | 532492 | 1182724 | 2420594 | 5182573 | 11434016 |
| **Pesticide metabolite** | | | | | | | | | |
| Hydroxy-isoproturon | 0.84 | 2047844 | 204714 | 322923 | 629194 | 1494965 | 5638538 | 29027773 | 141951008 |
| Dihydroxy-pyrimethanil | 0.72 | 565423 | 118901 | 202270 | 339479 | 486930 | 796518 | 2382533 | 14872204 |
| Hydroxy-simazine | 0.46 | 322421 | 90101 | 141837 | 215994 | 297794 | 418327 | 1100428 | 4460982 |
| Hydroxy-propamocarb | 0.38 | 3695460 | 245184 | 351372 | 1084626 | 3285529 | 8916329 | 76557394 | 161967056 |
| Fluazifop-desbuthyl | 0.19 | 1645847 | 231956 | 469280 | 899686 | 1352976 | 2408504 | 13507945 | 50272536 |
| Hydroxy-metazachlor | 0.18 | 392895 | 144754 | 173737 | 266947 | 372449 | 540294 | 1036031 | 3604622 |

Abbreviations. DR: detection rate. GM: geometric mean; Min: minimum value; Max: maximum value.

**Table S3.** Results of logistic regression models exploring the association between prenatal exposure levels ($log_2$-transformed) to pesticides and allergies in 6-year-old children (n = 387). Exposures with values BDL were imputed with the mean of 20 datasets generated using a truncated log-normal function.

| Compound ($log_2$ scaled) | Asthma | | | | Wheezing | | | | Eczema | | | |
|---|---|---|---|---|---|---|---|---|---|---|---|---|
| | Unadjusted | | Adjusted [a] | | Unadjusted | | Adjusted [a] | | Unadjusted | | Adjusted [a] | |
| | OR (95% CI) | *p*-value | OR (95% CI) | *p*-value | OR (95% CI) | *p*-value | OR (95% CI) | *p*-value | OR (95% CI) | *p*-value | OR (95% CI) | *p*-value |
| **Parent pesticide** | | | | | | | | | | | | |
| Metalaxyl | 0.85 (0.65 - 1.13) | 0.250 | 0.88 (0.67 - 1.17) | 0.356 | 1.02 (0.89 - 1.16) | 0.802 | 1.03 (0.91 - 1.18) | 0.626 | 0.88 (0.76 - 1.02) | 0.096 | 0.89 (0.77 - 1.04) | 0.146 |
| Carbetamide | 1.25 (0.93 - 1.70) | 0.145 | 1.24 (0.91 - 1.70) | 0.175 | 1.01 (0.88 - 1.14) | 0.940 | 1.00 (0.88 - 1.15) | 0.942 | 0.93 (0.80 - 1.08) | 0.357 | 0.92 (0.79 - 1.08) | 0.327 |
| Terbuthylazine | 0.98 (0.61 - 1.45) | 0.912 | 0.94 (0.58 - 1.42) | 0.798 | 1.17 (0.97 - 1.40) | 0.098 | 1.15 (0.96 - 1.39) | 0.137 | 0.85 (0.67 - 1.07) | 0.172 | 0.84 (0.66 - 1.06) | 0.159 |
| Imidacloprid | 1.07 (0.54 - 1.76) | 0.822 | 1.09 (0.54 - 1.85) | 0.775 | 0.97 (0.73 - 1.26) | 0.795 | 0.97 (0.74 - 1.27) | 0.824 | 0.79 (0.53 - 1.10) | 0.199 | 0.80 (0.54 - 1.13) | 0.240 |
| Flonicamid | 1.01 (0.77 - 1.25) | 0.901 | 0.98 (0.74 - 1.22) | 0.882 | 1.07 (0.96 - 1.18) | 0.202 | 1.05 (0.95 - 1.17) | 0.324 | 0.98 (0.85 - 1.10) | 0.706 | 0.95 (0.83 - 1.08) | 0.473 |
| **Pesticide metabolite** | | | | | | | | | | | | |
| Hydroxy-isoproturon | 0.85 (0.69 - 1.03) | 0.113 | 0.84 (0.68 - 1.02) | 0.095 | 0.95 (0.88 - 1.03) | 0.258 | 0.95 (0.87 - 1.03) | 0.232 | 1.02 (0.92 - 1.12) | 0.728 | 1.01 (0.92 - 1.12) | 0.791 |
| Dihydroxy-pyrimethanil | 1.35 (1.03 - 1.77) | 0.029 | **1.36 (1.04 - 1.80)** | **0.030** | 1.05 (0.94 - 1.18) | 0.385 | 1.05 (0.94 - 1.18) | 0.393 | 1.02 (0.89 - 1.18) | 0.731 | 1.02 (0.89 - 1.17) | 0.781 |
| Hydroxy-simazine | 0.95 (0.67 - 1.30) | 0.780 | 0.95 (0.66 - 1.33) | 0.793 | 1.05 (0.91 - 1.20) | 0.520 | 1.05 (0.91 - 1.21) | 0.526 | 0.94 (0.79 - 1.11) | 0.455 | 0.94 (0.78 - 1.11) | 0.450 |
| Hydroxy-propamocarb | 1.05 (0.90 - 1.21) | 0.490 | 1.10 (0.93 - 1.27) | 0.256 | 0.95 (0.88 - 1.02) | 0.131 | 0.95 (0.88 - 1.02) | 0.135 | 1.00 (0.92 - 1.09) | 0.957 | 1.01 (0.92 - 1.09) | 0.892 |
| Fluazifop-desbuthyl | 1.11 (0.84 - 1.39) | 0.387 | 1.15 (0.85 - 1.49) | 0.315 | 1.15 (1.01 - 1.31) | 0.031 | **1.15 (1.01 - 1.31)** | **0.036** | 1.00 (0.85 - 1.16) | 0.977 | 0.99 (0.84 - 1.15) | 0.936 |
| Hydroxy-metazachlor | 1.10 (0.64 - 1.64) | 0.686 | 1.12 (0.64 - 1.74) | 0.634 | 1.05 (0.85 - 1.31) | 0.640 | 1.06 (0.85 - 1.33) | 0.579 | 0.92 (0.68 - 1.19) | 0.530 | 0.93 (0.69 - 1.22) | 0.624 |

[a] Model adjusted for smoking/ETS exposure during pregnancy, breastfeeding up to 6 months, parental atopy history, parental education level, and child sex.

The odds ratios (ORs) in bold font are statistically significant: $p < 0.05$.

Abbreviations. BLD: below the detection limit.

**Table S4.** Output of logistic regression models examining the relationship between prenatal exposure levels ($log_2$-transformed) to pesticides and allergies in 6-year-old children (n = 387). Compounds with values BDL were imputed with LOD/√2.

| Compound ($log_2$ scaled) | Asthma | | | | Wheezing | | | | Eczema | | | |
|---|---|---|---|---|---|---|---|---|---|---|---|---|
| | Unadjusted | | Adjusted [a] | | Unadjusted | | Adjusted [a] | | Unadjusted | | Adjusted [a] | |
| | OR (95% CI) | *p*-value | OR (95% CI) | *p*-value | OR (95% CI) | *p*-value | OR (95% CI) | *p*-value | OR (95% CI) | *p*-value | OR (95% CI) | *p*-value |
| **Parent pesticide** | | | | | | | | | | | | |
| Metalaxyl | 0.84 (0.63 - 1.14) | 0.255 | 0.87 (0.65 - 1.18) | 0.363 | 1.02 (0.89 - 1.17) | 0.747 | 1.04 (0.91 - 1.19) | 0.572 | 0.88 (0.75 - 1.03) | 0.104 | 0.89 (0.76 - 1.05) | 0.169 |
| Carbetamide | 1.25 (0.91 - 1.72) | 0.164 | 1.24 (0.89 - 1.73) | 0.199 | 1.00 (0.87 - 1.15) | 0.968 | 1.00 (0.87 - 1.15) | 0.967 | 0.92 (0.78 - 1.09) | 0.336 | 0.92 (0.77 - 1.09) | 0.330 |
| Terbuthylazine | 0.97 (0.56 - 1.51) | 0.901 | 0.93 (0.54 - 1.47) | 0.784 | 1.20 (0.97 - 1.47) | 0.092 | 1.18 (0.95 - 1.46) | 0.131 | 0.82 (0.62 - 1.06) | 0.145 | 0.82 (0.61 - 1.06) | 0.146 |
| Imidacloprid | 1.09 (0.49 - 1.90) | 0.801 | 1.12 (0.50 - 2.02) | 0.733 | 0.96 (0.70 - 1.30) | 0.772 | 0.96 (0.70 - 1.32) | 0.818 | 0.71 (0.43 - 1.07) | 0.141 | 0.74 (0.45 - 1.11) | 0.183 |
| Flonicamid | 1.01 (0.76 - 1.26) | 0.916 | 0.98 (0.72 - 1.24) | 0.872 | 1.08 (0.96 - 1.20) | 0.187 | 1.06 (0.95 - 1.19) | 0.310 | 0.98 (0.85 - 1.11) | 0.730 | 0.95 (0.82 - 1.09) | 0.498 |
| **Pesticide metabolite** | | | | | | | | | | | | |
| Hydroxy-isoproturon | 0.83 (0.66 - 1.03) | 0.103 | 0.82 (0.65 - 1.02) | 0.091 | 0.96 (0.88 - 1.04) | 0.303 | 0.95 (0.87 - 1.04) | 0.276 | 1.02 (0.92 - 1.12) | 0.759 | 1.01 (0.91 - 1.12) | 0.806 |
| Dihydroxy-pyrimethanil | 1.35 (1.03 - 1.79) | 0.032 | **1.36 (1.03 - 1.82)** | **0.032** | 1.06 (0.93 - 1.20) | 0.397 | 1.06 (0.93 - 1.20) | 0.402 | 1.03 (0.88 - 1.19) | 0.738 | 1.02 (0.88 - 1.18) | 0.805 |
| Hydroxy-simazine | 0.95 (0.64 - 1.34) | 0.786 | 0.95 (0.63 - 1.37) | 0.788 | 1.05 (0.90 - 1.23) | 0.509 | 1.05 (0.90 - 1.23) | 0.519 | 0.93 (0.77 - 1.12) | 0.471 | 0.93 (0.77 - 1.12) | 0.460 |
| Hydroxy-propamocarb | 1.05 (0.88 - 1.23) | 0.545 | 1.10 (0.91 - 1.30) | 0.281 | 0.94 (0.87 - 1.02) | 0.125 | 0.94 (0.86 - 1.02) | 0.132 | 1.01 (0.91 - 1.10) | 0.912 | 1.01 (0.91 - 1.10) | 0.876 |
| Fluazifop-desbuthyl | 1.13 (0.83 - 1.43) | 0.388 | 1.17 (0.84 - 1.56) | 0.304 | 1.16 (1.01 - 1.35) | 0.037 | **1.16 (1.01 - 1.34)** | **0.043** | 1.00 (0.84 - 1.17) | 0.974 | 0.99 (0.82 - 1.16) | 0.868 |
| Hydroxy-metazachlor | 1.08 (0.58 - 1.70) | 0.759 | 1.11 (0.58 - 1.81) | 0.708 | 1.06 (0.83 - 1.35) | 0.631 | 1.08 (0.84 - 1.38) | 0.568 | 0.89 (0.63 - 1.19) | 0.462 | 0.90 (0.64 - 1.22) | 0.538 |

[a] Model adjusted for smoking/ETS exposure during pregnancy, breastfeeding up to 6 months, parental atopy history, parental education level, and child sex.

The odds ratios (ORs) in bold font are statistically significant: $p < 0.05$.

Abbreviations. BLD: below the detection limit. LOD: limit of detection.

### Supporting Figures

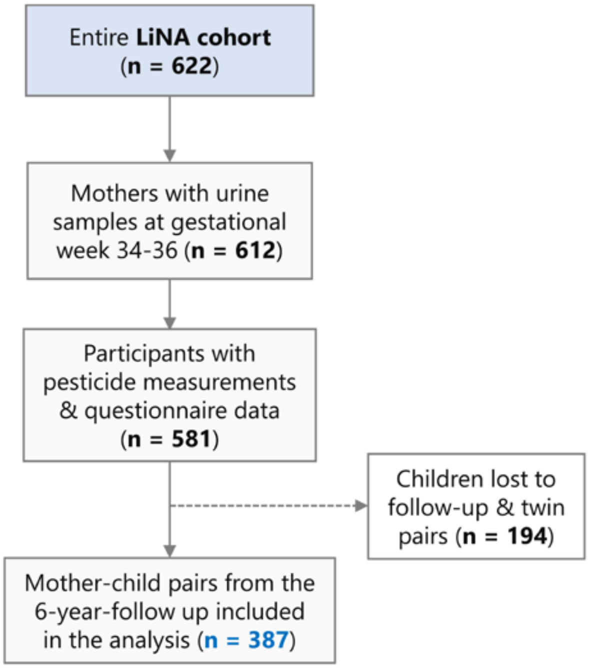


**Figure S1.** Flow chart of LiNA mothers (2006-2008) and paired 6-year-old children included in the study (n = 387).

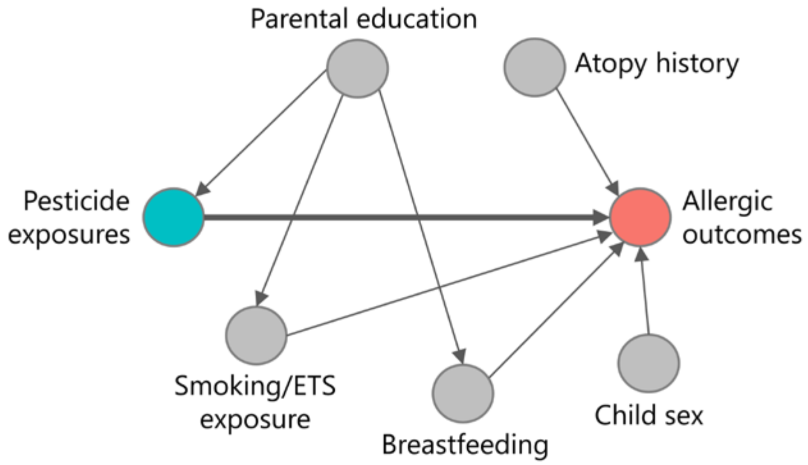


**Figure S2.** Directed acyclic graph (DAG) created to explore the causal pathway linking prenatal exposure to pesticides (light blue node) to allergic outcomes (red node), considering the effect of potential covariates (grey nodes). Regression models were adjusted for the covariates to estimate the total effect of the co-exposures on asthma, wheezing, and eczema.

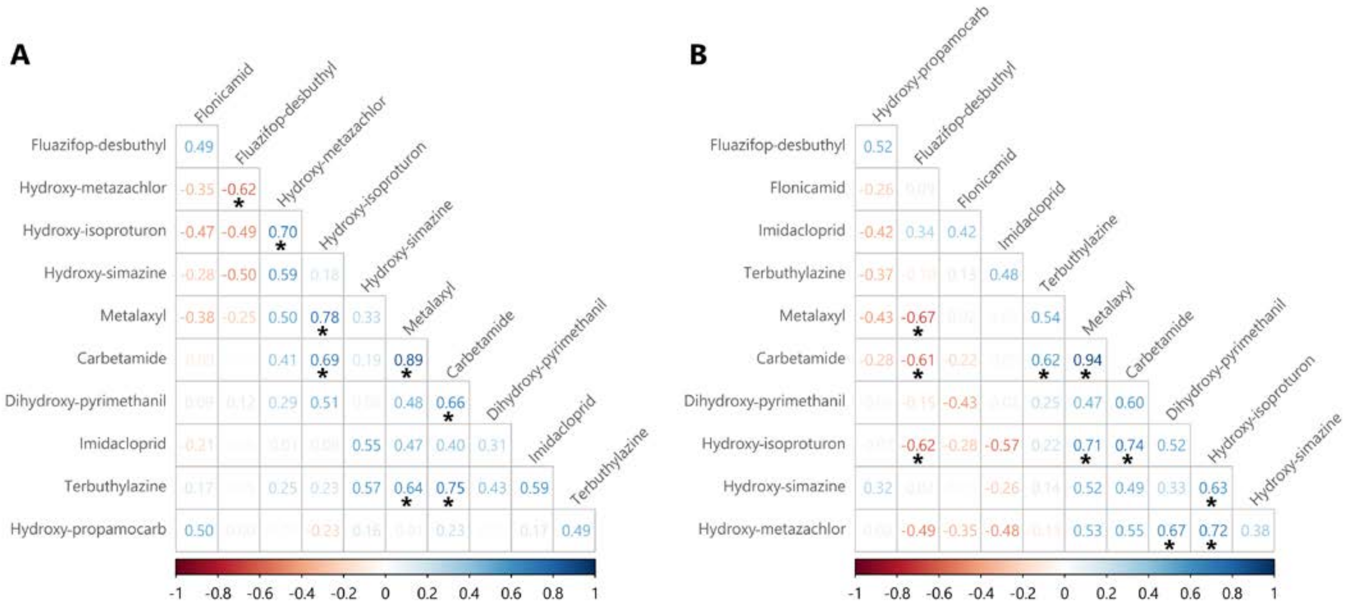


**Figure S3.** Pairwise Spearman's rank correlation matrices of chemical exposure levels in (**A**) the original dataset (LC-HRMS measurements) and (**B**) the LOD/√2-imputed dataset, where values BDL were replaced with the compound-specific minimum detection value divided by √2. In the color spectrum, blue and red shades show positive and negative correlations between the chemicals. The asterisk (*) denotes statistically significant correlations ($p < 0.05$).
